# Structural factors contributing to SARS-CoV-2 infection risk in the urban slum setting

**DOI:** 10.1101/2022.02.13.22270856

**Authors:** Mariam O. Fofana, Nivison Nery, Juan P. Aguilar Ticona, Emilia M.M.A. Belitardo, Renato Victoriano, Rôsangela O. Anjos, Moyra M. Portilho, Mayara C. de Santana, Laiara L. dos Santos, Daiana de Oliveira, Jaqueline S. Cruz, M. Cate Muencker, Ricardo Khouri, Elsio A. Wunder, Matthew D.T. Hitchings, Olatunji Johnson, Mitermayer G. Reis, Guilherme S. Ribeiro, Derek A.T. Cummings, Federico Costa, Albert I. Ko

## Abstract

**Background:** The structural environment of urban slums, including physical, demographic and socioeconomic attributes, renders inhabitants more vulnerable to SARS-CoV-2 infection. Yet, little is known about the specific determinants that contribute to high transmission within these communities.

**Methods and findings:** We performed a serosurvey of an established cohort of 2,035 urban slum residents from the city of Salvador, Brazil between November 2020 and February 2021, following the first COVID-19 pandemic wave in the country. We identified high SARS-CoV-2 seroprevalence (46.4%, 95% confidence interval [CI] 44.3-48.6%), particularly among female residents (48.7% [95% CI 45.9-51.6%] vs. 43.2% [95% CI 39.8-46.6%] among male residents), and among children (56.5% [95% CI 52.3-60.5%] vs. 42.4% [95% CI 39.9-45.0%] among adults). In multivariable models that accounted for household-level clustering, the odds ratio for SARS-CoV-2 seropositivity among children was 1.96 (95% CI 1.42-2.72) compared to adults aged 30-44 years. Adults residing in households with children were more likely to be seropositive; this effect was particularly prominent among individuals with age 30-44 and 60 years or more. Women living below the poverty threshold (daily per capita household income <$1.25) and those who were unemployed were more likely to be seropositive.

**Conclusions:** During a single wave of the COVID-19 pandemic, cumulative incidence as assessed by serology approached 50% in a Brazilian urban slum population. In contrast to observations from industrialized countries, SARS-CoV-2 incidence was highest among children, as well as women living in extreme poverty. These findings emphasize the need for targeted interventions that provide safe environments for children and mitigate the structural risks posed by crowding and poverty for the most vulnerable residents of urban slum communities.

## Introduction

More than 1 billion people who reside in urban slums or informal settlements are at increased risk of COVID-19, but also most likely to suffer loss of employment and income due to disease control measures such as lockdowns [1,2]. Although several studies have demonstrated an association of socioeconomic deprivation with increased COVID-19 risk and mortality, such studies have generally relied on ecological designs, or have compared poor communities to wealthier ones [3,4]. Moreover, studies relying on passive reporting of cases may suffer from surveillance bias. Prior studies of transmissible and non-transmissible diseases have shown variations in risk not only between communities but also within socially deprived environments, where certain segments of the population may be particularly vulnerable [5,6]. Yet, there remains little known about the SARS-CoV-2 risk gradients within urban slum communities and how these risk gradients may inform targeted interventions for this vulnerable population. Although the conditions and structure of each slum community are unique, there may be valuable common insights: regardless of geography, slums are invariably densely populated, have poor infrastructure, lack of access to services, and are inhabited by residents who experience significant housing and financial insecurity.

Brazil is among the countries that have suffered the highest burden of COVID-19, with more than 622,000 deaths reported as of February 2022 among its 213 million inhabitants over the course of several epidemic waves, the first of which occurred between May and September 2020 [7,8]. Brazil also has roughly 30 million people—16% of its urban population—residing in slum settlements within cities [9]. Prior to the pandemic, we had been performing long-term longitudinal follow-up of a cohort from the city of Salvador, Brazil to characterize the transmission dynamics and burden of leptospirosis [10], dengue [11], chiungunya [12] and Zika [13] among urban slum residents. Herein, we describe the findings of a serological survey of this cohort conducted after the first epidemic wave in Brazil in 2020, which aimed to examine the socioeconomic and structural factors that influence SARS-CoV-2 infection in the urban slum setting and characterize the gradients of risk within these deprived communities.

## Methods

### Study site and population

Our study was conducted in the Pau da Lima community in Salvador, Brazil, which has been previously described [10,11,13,14]. Geographically, Pau da Lima consists of hills and valleys, with a population of approximately 25,000 inhabitants per the most recent national census. The study site, shown in Fig. 1(A-C), encompasses four valleys, covering a densely populated area of 0.35 km^2^. Approximately half of the households do not hold legal titles to their homes, and more than 70% of the heads of household earn less than the Brazilian minimum wage [9]. As part of a longitudinal study started in 2001, individuals who reside in the community (defined as sleeping 3 nights or more per week in Pau da Lima), aged 2 years or more and who consented (parental consent for minors) were recruited into an open cohort and participated in household and serological surveys conducted annually or biannually. The most recent survey prior to the pandemic was conducted from September to November 2019 (Figure 1). A survey was performed between November 2020 and February 2021, after the first epidemic wave of COVID-19 and preceding the rollout of vaccines to the general population in February 2021, such that the presence of SARS-CoV-2 antibodies was reflective of prior infection.

**Figure 1.**
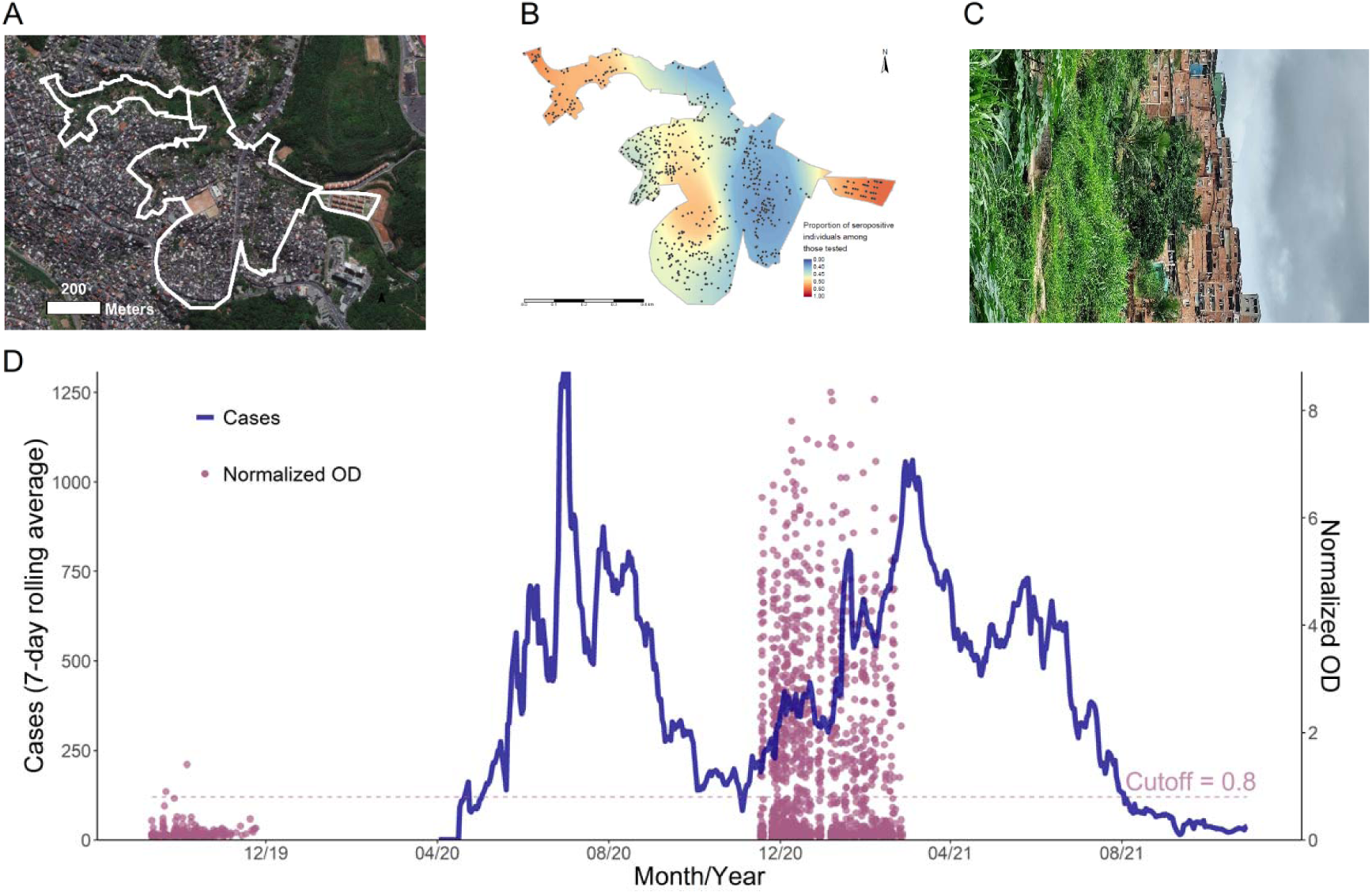
Study population and context. Panel A shows an aerial image of study area. Panel B depicts the location of participating households. The choropleth reflects the spatially adjusted seroprevalence within the study area. Panel C is a representative photo of the study area. D: Confirmed cases of COVID-19 in Salvador, Brazil, up to October 2021 (blue). Overlaid over the epidemiologic curve, in purple, are the normalized OD values of serological samples collected from cohort participants prior to the pandemic (September 11 to November 24, 2019) and during the second wave (November 17, 2020 to February 26, 2021).

### Data collection

Surveys and serum samples were collected between November 18, 2020 and February 26, 2021, after the first epidemic wave and in the initial stages of the second wave (Fig. 1D). A community field research team conducted a census of each household, listing the number of residents, gender, and number of people per room. The head of household contributed information on the income of each wage-earning resident in the household. Household income per capita was estimated as the total income of all residents divided by the number of residents in each household. Participants who consented also completed a detailed individual-level survey, including data on COVID-19 exposures, COVID-19 cases in the household, preventive behaviors (e.g. handwashing, mask use), medical history, and symptom history. The survey includes participant-reported socioeconomic variables such as type of employment, race/ethnicity, and level of education. Blood samples were collected by venipuncture from consenting participants and stored in a cooler for transport to the laboratory facility. After centrifugation, the obtained sera were aliquoted and maintained at -20°C until analysis.

### Laboratory methods

We measured anti-S IgG in serum samples collected from all cohort participants after the first epidemic wave, and from a subset of participants in a survey conducted prior to the introduction of SARS-CoV-2 in Brazil. Levels of immunoglobulin G against the SARS-CoV-2 spike protein (anti-S IgG) were measured using a commercially available ELISA kit (Euroimmun AG, Lübeck, Germany) according to the manufacturer’s instructions. Samples were diluted 1:100 in sample buffer, and a set of calibrator, positive, and negative control samples were included on each plate. Optical density (OD) was measured at a wavelength of 450 nm and normalized values were derived by dividing the OD value of each test sample by the value of the calibrator. All samples with a normalized OD ≥ 0.8 were defined as positive, based on a Bayesian estimation of optimal cutoffs (Fig S2). We also measured anti-S IgG in a subset of serum samples collected from cohort participants prior to the introduction of SARS-CoV-2 in Brazil.

### Data analysis

The primary outcome was SARS-CoV-2 seropositivity as assessed by anti-S IgG. Independent variables included demographic, socioeconomic, and household variables as assessed from the household and individual surveys. For categorical variables, proportions were compared using the Chi-square test. For continuous variables, we first conducted a visual assessment of correlation with SARS-CoV-2 anti-S IgG positivity and compared distributions using the Wilcoxon rank-sum test. We assessed measures of association using univariable logistic regression. For variables that appeared to have a non-linear relationship with seropositivity risk, we used a spline in a generalized additive model. Variables with a statistically significant (p<0.05) association with SARS-CoV-2 seropositivity were included in a multivariable logistic regression model.

Our descriptive analyses revealed higher seroprevalence among participants aged <18 years compared to adults. To further elucidate (1) whether children are more susceptible to COVID-19 and (2) whether they contribute to higher transmission in this setting, we performed a multilevel, multivariable logistic regression that incorporated clustering at the household level and nonlinear effects of age. Individual and per capita income both showed a strong positive correlation with age among adults; we therefore did not include income and age simultaneously in the model. We assessed model fit based on Akaike Information Criterion (AIC) to select the most parsimonious explanatory model.

As we noted a higher seroprevalence among women compared to men, we conducted a similar analysis among the subset of adult participants (aged ≥18 years), to assess whether the association between gender and SARS-CoV-2 exposure was mediated by income, employment, and household structure in this population. Participants with missing values for independent variables were excluded from the relevant analyses (pairwise deletion). All analyses were conducted using the software R, version 4.1.1 [15].

### Ethical considerations

The study was approved by the Institutional Review Boards of the Gonçalo Moniz Institute, Oswaldo Cruz Foundation (Fiocruz) and the Brazilian National Commission for Ethics in Research (CAAE 35405320.0.1001.5030 and 17963519.0.0000.0040), and the Yale University Human Research Protection Program (2000031554).

## Results

### Population demographics

A total of 2,035 individuals (1,456 adults and 579 children) in 948 households were recruited from a total of 2,481eligible residents (Figure S1). Serological samples were obtained for every resident in 403 households (917 individuals). Table 1 describes the demographic and socioeconomic characteristics of the study population. The median age was 29 years [IQR 16-44] and 58% were female. The median size of participating households was 3 [IQR 2-4], with a median household daily per capita income of 2.32 USD [IQR 0.80-4.64]. Most participants reported their ethnicity as Black (51%) or Brown (42%). Overall, 35% (500/1,427) of adult participants had six or fewer years of formal education, and 58% (748/1,288) were unemployed. The majority of participants reported frequent (every day or most days) adherence to handwashing (70.5%), alcohol gel use (67.0%), and face mask use (72.5%), while fewer participants reported frequent social distancing (43.4%) or physical distancing of 2 meters in public spaces (46.6%).

**Table 1:**
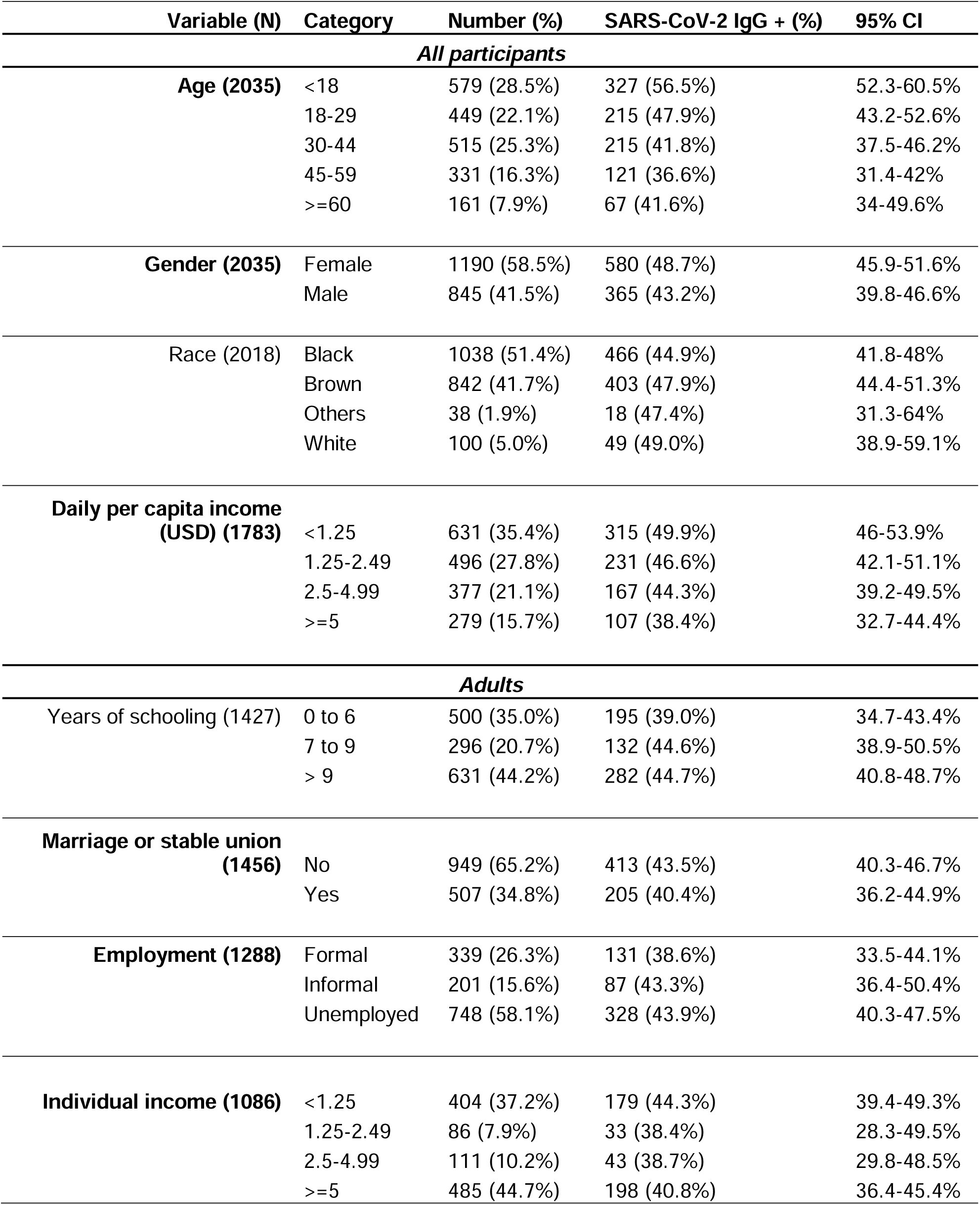
Study population and demographic characteristics. Employment, education and marriage were assessed for adult (>18 years) participants. Variables with statistically significant effects in univariable analyses are indicated in bold.

### Seroprevalence

Figure 1D shows the serology assay results for the cohort, overlaid on the epidemic curve of confirmed COVID-19 cases in Salvador, Brazil. Among 195 samples that were selected and tested from the pre-pandemic survey, September 9 to November 11, 2019, two (1.0%) were positive. Fitting the observed anti-S IgG levels with a Bayesian mixture model revealed little overlap between the predicted distributions of OD values for individuals with and without a serological response (Figure S2). Based on this model, a cutoff of 0.8 has greater than 95% specificity in identifying serological response (Table S1).

Of the 2,035 samples collected from November 2020 to February 2021, a total of 945 (46.4%, 95% confidence interval [CI] 44.3-48.6%) had positive IgG levels against SARS-CoV-2. Among the 945 seropositive individuals, 273 (28.9% [95% CI 26.0-31.9%]) reported having at least one COVID-19 related symptom (cough, coryza, sore throat, shortness of breath, fever, chills, anosmia, dysgeusia or headache) since the beginning of the pandemic. The frequency of reported symptoms was lower among seronegative individuals (19.4% [95% CI 17.1-21.9%]; difference 9.5% [95% CI 5.7-13.4%]). The proportion of households with at least one seropositive individual was 62.3%.

### Risk factors

Seroprevalence was significantly higher among children (56.5% [95% CI 52.3-60.5%]) than adults (42.4% [39.9-45.0%]) and in female (48.7% [45.9-51.6%]) compared to male (43.2% [39.8-46.6%]) participants (Table 1, Figures 2A and 2B). Additional variables associated with seropositivity in univariable analyses included low per capita household income and residents per household among all participants, and unemployment and not being married or in a stable union among adults. We did not observe a significant association with race or education level (Tables 1 and S2). The reported use of non-pharmaceutical interventions was not significantly associated with the risk of seropositivity (Table S3). Because risk factors (including structural environment of the home, health attitudes, and preventive behaviors) are likely to be correlated among members of a household, we further conducted an analysis using a multilevel multivariable model to account for household-level clustering of risk.

**Figure 2.**
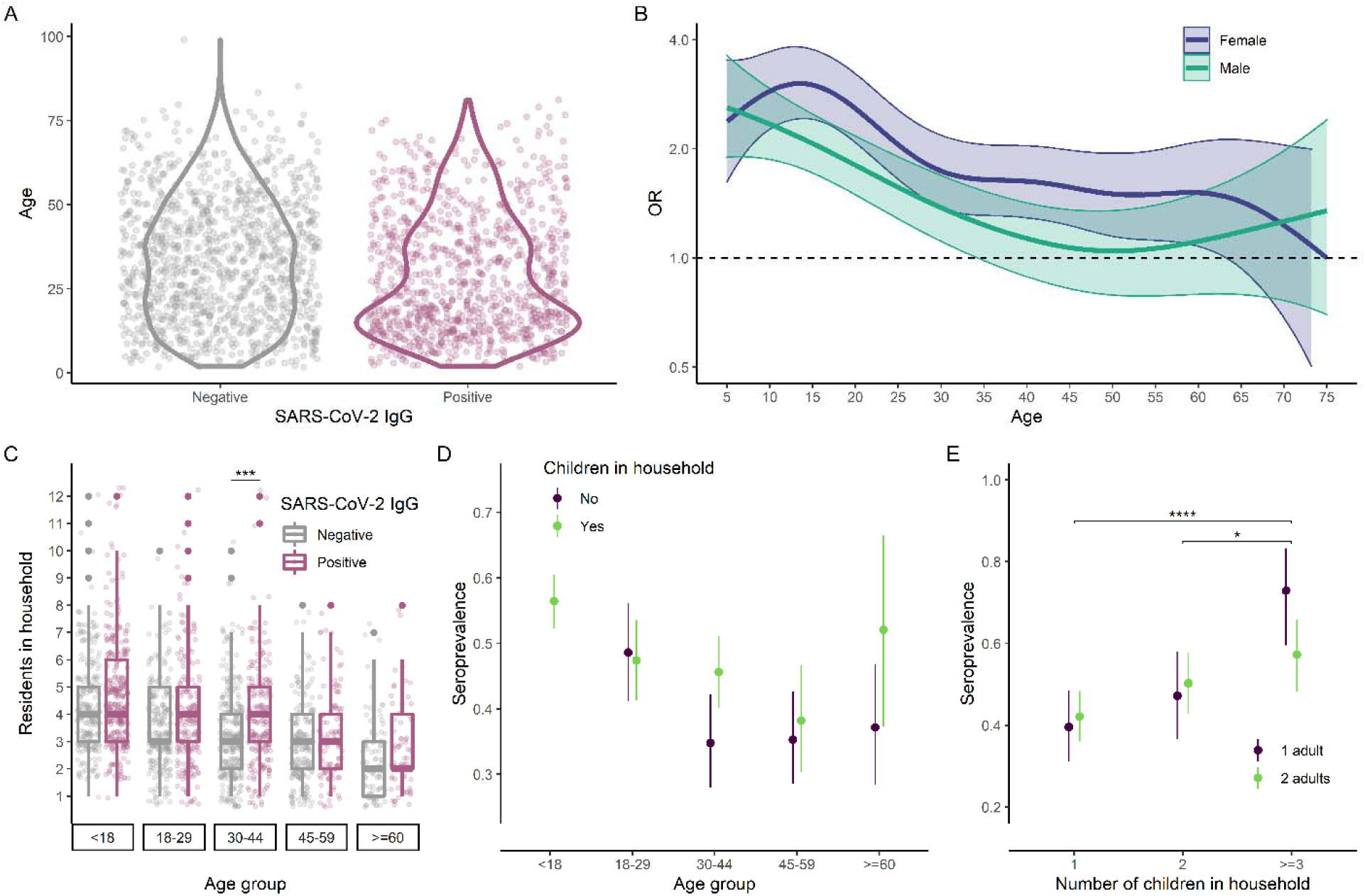
Seroprevalence among children and in their households. A: Age distribution of SARS-CoV-2 seronegative (median 33, interquartile range [IQR] 19-46) and seropositive (median 24 years, IQR 14.0-41) individuals. B: Odds ratio (OR) for SARS-CoV-2 seropositivity and 95% confidence interval (CI) associated with age, as estimated using a generalized additive model. C: Distribution of household size by age among SARS-CoV-2 seronegative and seropositive individuals. Seropositive individuals tended to be in larger households compared to seronegative individuals in the same age group. D: OR and 95% CI stratified by age group and presence of children in the household. E: Variation in seroprevalence among children by household composition (number of children and number of adults). The number of other children in the household was associated with higher seroprevalence, but there was no statistically significant difference between households with one adult and those with two adults. Asterisks indicate statistically significant differences (Bonferroni-adjusted p<0.05: *; <0.01: **; <0.001: ***).

### Household composition

Adults who have larger households, and those who shared their residence with children were more likely to be SARS-CoV-2 seropositive (Fig 2C and 2D). The effect of the presence of children was particularly pronounced among adults aged 30-44 years (OR 1.57, 95% CI 1.08-2.28) and those aged 60 years or more (OR 1.84, 95% CI 0.93-3.64). Children were also more likely to be seropositive if they lived with other children, but there was no significant difference in seroprevalence between children who lived with one adult and those who lived with two adults (Fig 2E).

To examine whether the higher seroprevalence among children was mediated by increased crowding in households with children, we implemented binomial regression models with random intercepts per household to account for clustering, and compared several alternative models (Table 2). First, we compared a baseline model including a binary variable for the presence of children in the household (Table 2, Model 1) to an alternative model including the total number of residents in the household (Model 2). Model 2 demonstrated a better fit based on the AIC. We further conducted a mediation analysis on the effect of the presence of children in the household. The OR for the association of presence of children with SARS-CoV-2 seropositivity in Model 1 was 1.32 (95% CI 0.94-1.87). However, after including household size as a variable in Model 3, the effect measure was attenuated (OR 0.90, 95% CI 0.61-1.32), consistent with a mediation effect. We observed similar findings in models that included income rather than age (Table S5). In the three main models, the OR for SARS-CoV-2 seropositivity among children remained stable (2.08 [1.49-2.89] to 1.96 [1.42-2.72]), suggesting that the higher seropositivity among children was not entirely attributable to household size.

**Table 2:** Effect of children in household on odds of SARS-CoV-2 seropositivity. All models include age (A) and gender (G) as independent variables. Additionally, Model 1 includes a binary variable for the presence of children in the household (C), whereas Model 2 includes the total number of residents in the household (R). Model 3 includes all of the above variables.

|  |  | **Model 1**<br>**(AGC)** | **Model 2**<br>**(AGR)** | **Model 3**<br>**(AGCR)** |
| --- | --- | --- | --- | --- |
| **Variable** | **Category** |  |  |  |
| **Age** | <18 | 2.08 (1.49-2.89) | 1.96 (1.42-2.72) | 1.99 (1.43-2.77) |
|  | 18-29 | 1.44 (1.02-2.05) | 1.39 (0.98-1.98) | 1.38 (0.97-1.97) |
|  | 30-44 | (Ref) | (Ref) | (Ref) |
|  | 45-59 | 0.74 (0.5-1.11) | 0.77 (0.52-1.14) | 0.76 (0.51-1.13) |
|  | >=60 | 1.13 (0.67-1.91) | 1.24 (0.74-2.08) | 1.22 (0.72-2.05) |
| **Gender** | Male | (Ref) | (Ref) | (Ref) |
|  | Female | 1.53 (1.2-1.95) | 1.50 (1.18-1.91) | 1.51 (1.18-1.92) |
| **Children in home**<br>**(Y/N)** |  | 1.32 (0.94-1.87) |  | 0.90 (0.61-1.32) |
| **Total residents** |  |  | 1.23 (1.12-1.35) | 1.24 (1.12-1.38) |
| **AIC** |  | 2625 | 2608 | 2610 |
| **ICC** |  | 0.408 | 0.397 | 0.397 |

### Poverty and unemployment among women

We explored the associations between gender and sociodemographic characteristics to elucidate the mechanisms underlying the significantly higher SARS-CoV-2 positivity in women compared to men (48.7% vs 43.2%, p = 0.015). Women were more likely to be unemployed than men (68.0% [95% CI 64.7-71.2%] vs 41.5% [95% CI 37.1-46.1%]) and had lower income per capita in their households (Fig 3A) regardless of employment status (median [interquartile range] 2.44 [1.56-4.64] vs 3.41 [1.78-6.67] USD/day among employed adults; 1.60 [0-3.33] vs 1.74 [0-4.00] USD/day among unemployed adults). After accounting for income and employment, women had increased risk for SARS-CoV-2 seropositivity compared to men (Fig 3B). Women who were unemployed and had a household per capita income below 1.25 US dollars per day were significantly more likely to be seropositive compared to men with the same employment and income status (OR 2.5, 95% CI 1.9-3.4; Fig 3B).

**Figure 3.**
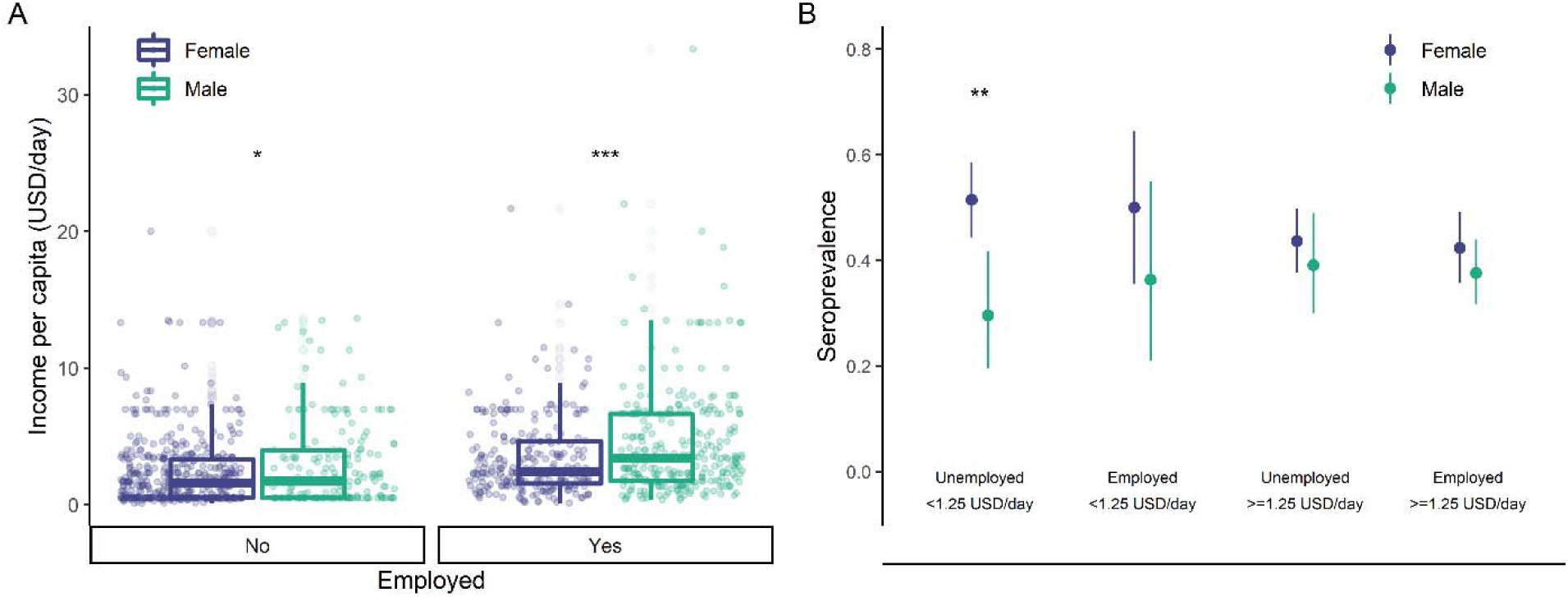
Socioeconomic vulnerability among women. A: Household income (per capita) by employment status and gender. B: Seroprevalence by household income (per capita), employment status and gender. Asterisks indicate statistically significant differences (Bonferroni-adjusted p<0.05: *; <0.01: **; <0.001: ***).

## Discussion

From the early days of the COVID-19 pandemic, there was recognition that the fragile structural and social environment of urban slums constitutes an important yet poorly understood risk [2,16]. Although studies have described socioeconomic disparities in COVID-19 risk and outcomes [17,18], few have focused on the particular risk of urban slums. In this community-based study conducted in the city of Salvador, Brazil, we found that nearly half of residents in an urban slum had evidence of SARS-CoV-2 infection after the first wave of the epidemic in 2020. In contrast, a national, regionally stratified serosurvey conducted in June 2020 estimated seroprevalence to be 5.5% in Salvador and 3.2% in the Northeast region where the city is situated [19]. This finding is consistent with studies in other settings that reported higher seroprevalence in slums than in non-slum settings [4,20], highlighting the increased vulnerability of socioeconomically deprived communities.

We were able to collect detailed demographic and socioeconomic data at both individual and household levels and overcome the limitations of previous studies. Although a few studies have investigated COVID-19 risk in slum compared to non-slum areas, none to date has examined the gradients of risk within a slum community. Our findings suggest that even within an overall socioeconomically deprived environment, there is a gradient of risk associated with income, employment, and household composition.

We found that children in this urban slum setting had high SARS-CoV-2 seroprevalence, significantly greater than observed in adults, which sharply contrasts with patterns of transmission in high-income countries and urban populations within these countries. Most studies to date have reported lower or similar seroprevalence of COVID-19 among children compared to adults during the early phase of the pandemic [21]. Serosurveys relying on residual clinical samples have reported higher prevalence among children, but may not be reflective of the general population of children who do not regularly undergo blood draws [22–24]. The community-based design of our study allows for better comparability of seroprevalence between children and adults.

Prior studies of respiratory viruses such as influenza showed that school-aged children are reservoirs of transmission [25,26] but it remains unclear whether this is the case with SARS-CoV-2, particularly in densely populated and socially deprived environments such as urban slums. Interestingly, we found that adults aged 30-44 and 60 years or more were more likely to be SARS-CoV-2 seropositive if they lived with children, suggesting that multigenerational composition of urban slum households contributes to infection risk in specific adult groups (age bearing parents and grandparents). Yet, we were unable due to the cross-sectional design to determine whether children were more likely to be the index case within households.

Although SARS-CoV-2 seropositivity was associated with larger household size; however, the ORs for children were similar in models that did and did not include the total number of household residents, suggesting that the high seropositivity in children cannot be exclusively attributed to household size. Schools in Salvador closed in-person instruction for a prolonged period from March 2020 to May 2021 [27]. Yet, children may have continued to socialize in similarly assortative patterns within their urban slum community (e.g., contact with other children in neighboring houses). The combination of school closures and the lack of safe childcare options and targeted public health prevention may have resulted in higher exposure to SARS-CoV-2 in high-risk urban slum environments, as suggested by the high incidence during a single pandemic wave in this study.

We found that women in this urban slum community had significantly higher risk compared to men. Although there is evidence for sex differences in the immune response to SARS-CoV-2 infection and disease severity [28], serological surveys have generally found similar or lower prevalence among women compared to men [29]. Thus, the gender difference in risk observed in our study was likely driven by social rather than biological determinants. Interestingly higher seroprevalence among women has also been reported in other urban slum settings: two serosurveys conducted in urban slums Mumbai and Bangalore, India, reported a higher seroprevalence among women than men, but did not investigate potential mediators of this effect [4,30]. We found that women who were unemployed and had the lowest household income were significantly more likely to be seropositive for SARS-CoV-2. Thus, in addition to the disparity between slum and non-slum areas, the risk gradient within slums results in even higher SARS-CoV-2 infection risk for the poorest women.

A limitation of this study is that our data do not allow for identification of time of infection nor of primary cases within households, such that we were unable identify which household members were first to be infected and estimate what proportion of SARS-CoV-2 transmission occurred within households. While the majority of the survey occurred during the nadir between two waves, some participants were surveyed during the initial phase of the second wave. Seroprevalence may thus overestimate infection risk attributable to the first wave. Our observational design does not allow for a complete decomposition of the complex interactions between gender, employment, income, household structure and other unmeasured variables: households with children are larger, and larger households have lower per capita income, such that regression model estimates are not truly independent. Nevertheless, our data highlight the need for targeted interventions that address the risk experienced by the most vulnerable segments of slum communities, such as women who are unemployed, and the poorest households.

Potential sources of bias include selection bias as participants in the study may differ from residents who chose not to participate. Ascertainment and recall bias cannot be excluded, as many of the exposure variables were participant-reported. Finally, we did not have complete serological data for all household members. However, in sensitivity analyses using only data from households with serological data for all residents, our findings remained consistent (Figures S4, S5, S6).

### Conclusions

Our study provides key insights into the gradient of SARS-CoV-2 infection risk within the deprived environment of urban slums, leveraging a large community-based cohort with detailed demographic and socioeconomic data at both individual and household levels. While the specific micro-environment of each community is unique, Pau da Lima shares features with urban slum communities in general: vulnerability to COVID-19 derives from the physical environment (construction quality, crowding, poor ventilation, poor access to sanitation facilities), as well as the social environment (financial precarity, mobility, social contact patterns) [31,32]. For example, slum residents are less able to isolate as they must travel to work and maintain an income and depend on often crowded and poorly ventilated public transportation to places of work typically located far from slum areas [33].

As has become evident, effective responses to the COVID-19 pandemic must include not only biomedical interventions but also the deployment of social support systems to mitigate the profound disruptions caused by the disease itself and control measures [34,35]. Participants in our study reported high adherence to hand hygiene and mask use but less frequent social and physical distancing, possibly because such measures are less feasible and effective in the crowded environment of slums. Moreover, given that the structural risk factors that we have identified cannot be readily modified, interventions such as vaccination that can mitigate risk independent of socioeconomic status remain crucial. Despite concerns about vaccine hesitancy, we previously reported 66% acceptance of vaccination for SARS-CoV-2 among adults in this community, similar to countries such as the United States [14,36]. Bridging the gap in vaccine access for populations in low- and middle-income countries, where most urban slums are located, must be a priority to achieve an equitable pandemic response.

## Data Availability

All data produced in the present study are available upon reasonable request to the authors

