## Supplemental materials for "Structural factors contributing to SARS-CoV-2 infection risk in the urban slum setting"

**Supplementary materials**

*Study flowchart*

A detailed census of households in the study area, including number and demographic characteristics of residents in each household, was performed to identify eligible individuals. Individuals who consented to participated in the study underwent an individual survey as well as collection of a serum sample to measure SARS-CoV-2 antibody levels (Figure S1).

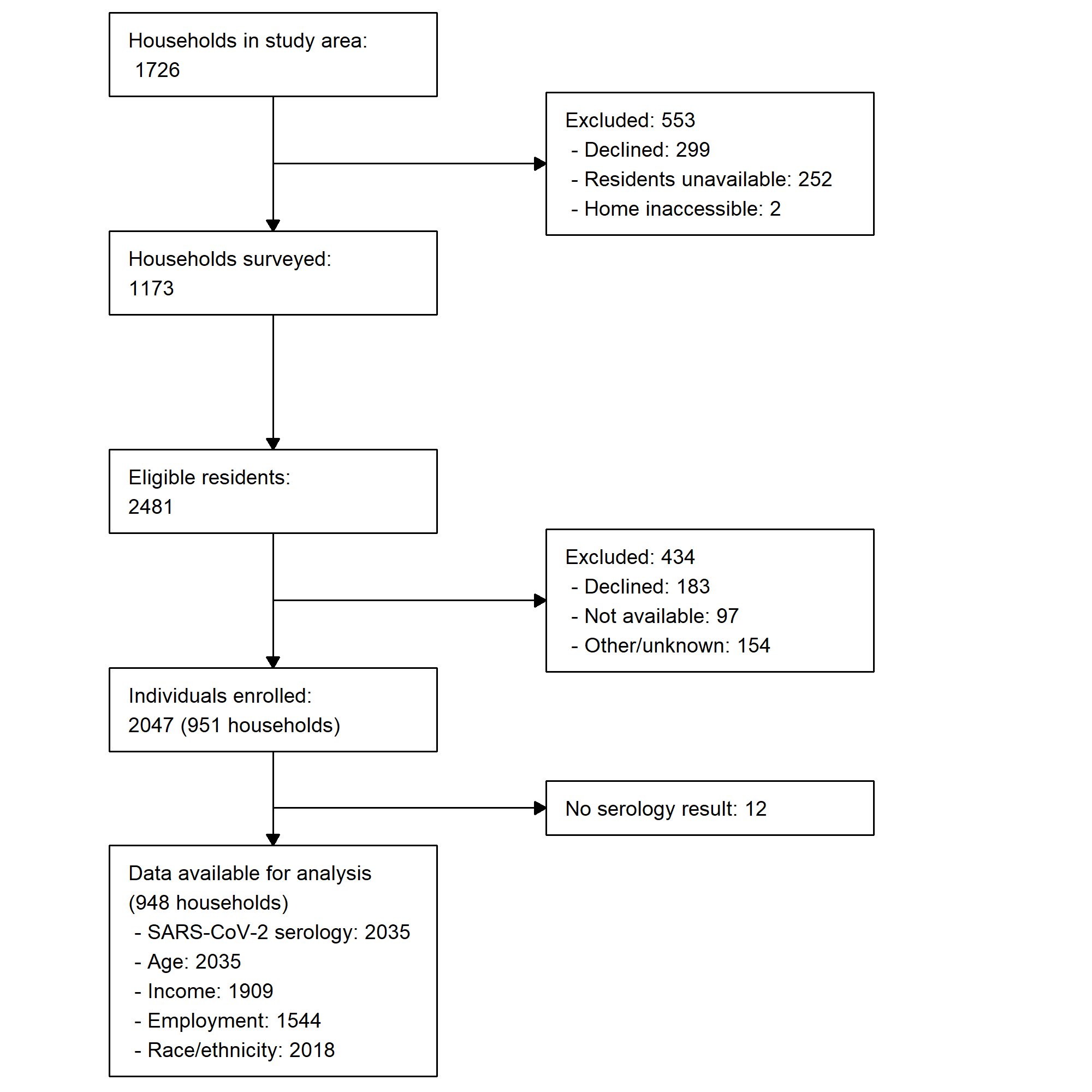

**Figure S1**: Study eligibility and recruitment flowchart

*Distribution of normalized optical density values*

The distribution of OD ratio values in our sample suggests that there is overlap in antibody levels between patients with and without SARS-CoV-2 infection, as observed in prior studies [1,2]. Per the manufacturer, certain test values are considered borderline, but it is unclear whether such results are more likely to represent infected or non-infected patients. We therefore conducted a sensitivity analysis, considering alternate cutoff values obtained from a Bayesian mixture model, an extension of Hitchings et al [3].

The model assumes that there are two separate populations: participants with and without SARS-CoV-2 infection. Given proposed prior distributions for the OD values for these two populations, we evaluate the likelihood of the observed data and use a MCMC algorithm to yield posterior distributions. We assumed normalized OD values fit a mixture of two Gaussian distributions. We based the prior distribution for individuals without serological response (non-responders) on the observed values from samples collected prior to the COVID-19 pandemic (mean 0.141, SD 0.143). The prior parameters for normalized OD values among individuals with serological response were drawn from uniform distributions.

We then implemented an MCMC algorithms with a chain length of 250,000, a burn-in of 125,000 simulations and an acceptance ratio of 0.2. The likelihood was computed as follows:

$$l\left( \theta\right)= \sum_{i} log\left( p*f_{R}\left( x_{i} \right)+\left( 1-p \right)*f_{NR}\left( x_{i} \right) \right)$$

where *p* is the probability of being a responder, *f_R_(x)* is the pdf of the seroresponse distribution, and *f_NR_(x)* is the pdf of the non-response distribution.

Figure S2A shows the observed (black line) density of normalized OD values and the posterior distributions obtained from the Bayesian mixture model (non-responders in red, responders in green, combined distribution in blue). Table S1 shows the specificity associated with various OD thresholds to identify individuals with a positive serological response. For a specificity of 99.5%, the model-derived cutoff is 1.05, similar to the manufacturer-recommended cutoff of 1.1 (Figure S2B). When stratifying by age group, the model-derived cutoff was slightly higher among participants 15 years and younger compared to older participants (Figure S2C), but the cutoff remained within the manufacturer-suggested range. In order to confirm that the observed higher seroprevalence among women was not due to differences in antibody response (i.e., women having higher antibody levels and thus being more likely to be identified as seropositive), we compared the distribution of normalized OD values (Figure S2D). A difference in antibody response would be reflected by a shift in the density curves on the x-axis, which we did not observe.

**Table S1: Seropositivity thresholds**

| **Normalized OD threshold** | **Specificity** |
| --- | --- |
| 0.765 | 95.0% |
| 0.869 | 98.0% |
| 0.955 | 99.0% |
| 1.05 | 99.5% |
| 1.295 | 99.9% |

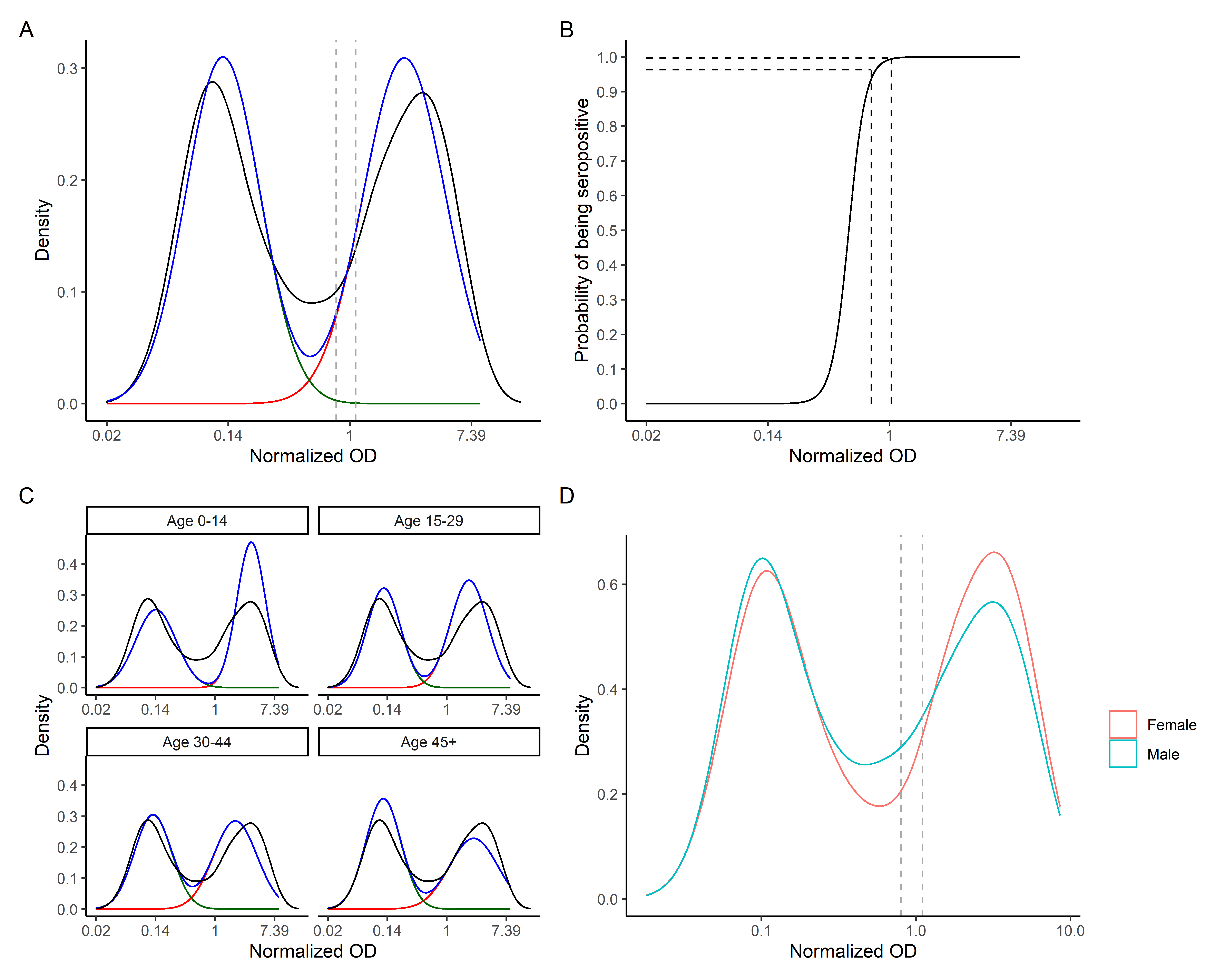
**Figure S2**: Observed and fitted distributions of normalized OD values. Panels A and C show the observed distribution of normalized OD values (black lines) for the overall study population (A) and by age group (C). Fitted distributions obtained from the MCMC mixture model are shown for seropositive (red line), seronegative (green line), and all (blue line) individuals. Panel B shows the model-predicted probability of SARS-CoV-2 seropositivity. Panel D shows the distribution of normalized OD values among male and female participants. Dashed lines (panels A, B, D) represent the manufacturer-recommended cutoffs of 0.8 and 1.1 for the anti-S1 IgG ELISA.

*Household composition and increased risk among women*

In addition to differences in employment and income, there were differences between adult women and men in their household composition. Compared to men, women more often lived in larger households and with children. Moreover, women were more likely than men to be the single adult in a household with children (Figure S3). Women with larger households dependent on a single income were thus more likely to experience risk from both physical crowding and poverty.

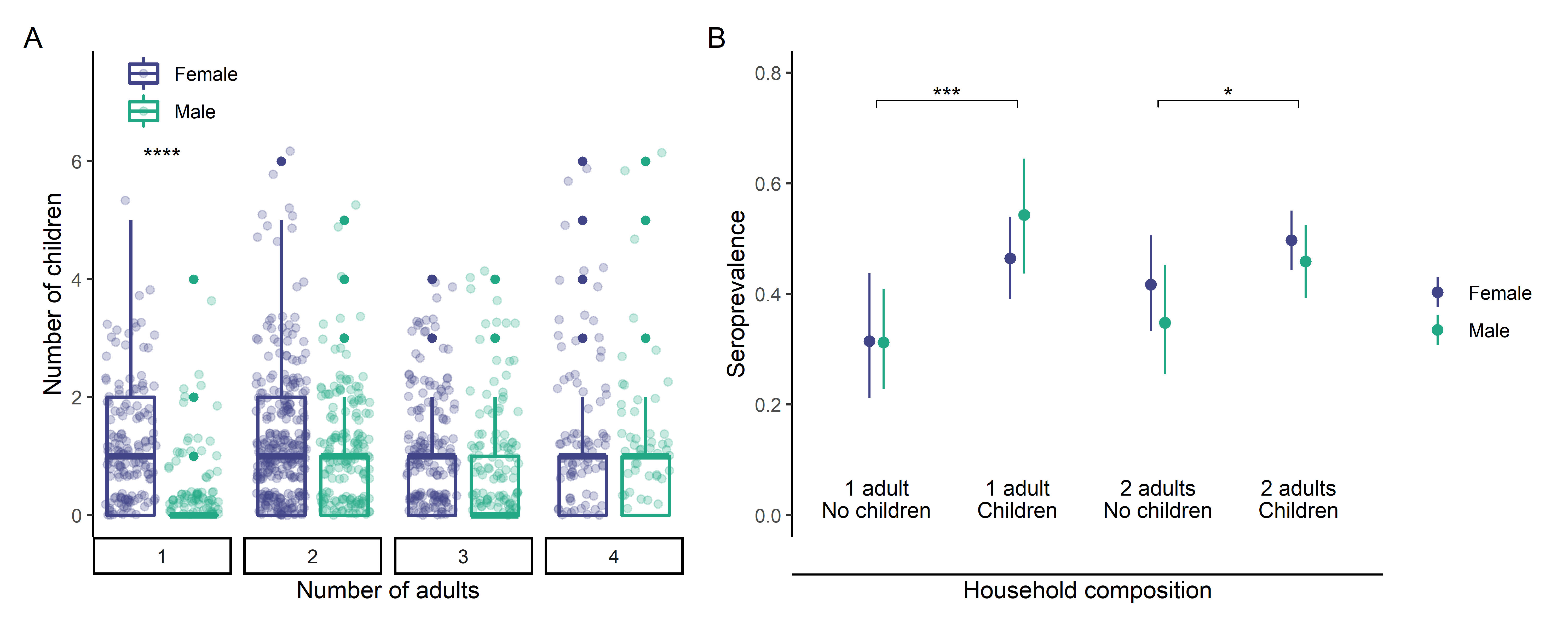
**Figure S3**. A: Household composition by gender. Women were more likely to be the sole adult in households with multiple children. B: Variation in seroprevalence among adults by household composition (number of adults and presence of children). Adults living with children were more likely to be seropositive. Accounting for household composition, there was no statistically significant difference in seroprevalence between men and women.

**Table S2: Univariable analyses**

| **Variable (N)** | **Category** | **OR (95% CI)** |
| --- | --- | --- |
| Age | <18 | **2.31 (1.57-3.38)** |
|  | 18-29 | **1.38 (1.02-1.87)** |
|  | 30-44 | (Ref) |
|  | 45-59 | 0.99 (0.7-1.38) |
|  | >=60 | 1.25 (0.79-1.95) |
| Gender | Male | (Ref) |
|  | Female | **1.43 (1.13-1.8)** |
| Race | Black | (Ref) |
|  | Brown | 1.13 (0.9-1.41) |
|  | White | 1.14 (0.67-1.92) |
|  | Other | 0.84 (0.26-2.68) |
| Marriage or stable union | No | (Ref) |
|  | Yes | 0.91 (0.7-1.17) |
| Years of schooling | 0 to 6 | 0.87 (0.65-1.17) |
|  | 7 to 9 | 0.87 (0.64-1.17) |
|  | >9 | (Ref) |
| Employment | Formal | (Ref) |
|  | Informal | 1.29 (0.89-1.85) |
|  | Unemployed | 1.05 (0.8-1.38) |
| Daily per capita income | <1.25 USD/day | 1.19 (0.81-1.74) |
|  | 1.25-2.49 USD/day | 1.06 (0.74-1.53) |
|  | 2.5-4.99 USD/day | 1.13 (0.79-1.62) |
|  | ≥5 USD/day | (Ref) |
| Total residents in household | Per person | **1.15 (1.07-1.24)** |

**Table S3: Adherence to non-pharmaceutical interventions**

| **Variable (N)** | **Frequency (per week)** | **Number (%)** | **SARS-CoV-2 IgG + (%)** | **95% CI** |
| --- | --- | --- | --- | --- |
| Handwashing (2032) | Never | 135 (6.6%) | 62 (45.9%) | 37.4-54.7% |
|  | A few days | 464 (22.8%) | 224 (48.3%) | 43.7-52.9% |
|  | Most days | 308 (15.2%) | 158 (51.3%) | 45.6-57.0% |
|  | Every day | 1125 (55.4%) | 499 (44.4%) | 41.4-47.3% |
| Alcohol-based sanitizer use (2032) | Never | 174 (8.6%) | 72 (41.4%) | 34.1-49.1% |
|  | A few days | 496 (24.4%) | 240 (48.4%) | 43.9-52.9% |
|  | Most days | 302 (14.9%) | 151 (50.0%) | 44.4-55.6% |
|  | Every day | 1060 (52.2%) | 480 (45.3%) | 42.3-48.3% |
| Mask use (2022) | Never | 84 (4.2%) | 33 (39.3%) | 29.0-50.6% |
|  | A few days | 472 (23.3%) | 231 (48.9%) | 44.4-53.5% |
|  | Most days | 309 (15.3%) | 150 (48.5%) | 42.9-54.3% |
|  | Every day | 1157 (57.2%) | 524 (45.3%) | 42.4-48.2% |
| Physical distancing >2m (2029) | Never | 383 (18.9%) | 179 (46.7%) | 41.7-51.9% |
|  | A few days | 700 (34.5%) | 336 (48.0%) | 44.2-51.8% |
|  | Most days | 385 (19.0%) | 187 (48.6%) | 43.5-53.7% |
|  | Every day | 561 (27.6%) | 240 (42.8%) | 38.7-47.0% |
| Isolation (2031) | Never | 426 (21.0%) | 196 (46.0%) | 41.2-50.9% |
|  | A few days | 724 (35.6%) | 354 (48.9%) | 45.2-52.6% |
|  | Most days | 390 (19.2%) | 171 (43.8%) | 38.9-48.9% |
|  | Every day | 491 (24.2%) | 222 (45.2%) | 40.8-49.7% |

*Sensitivity analyses*

A sensitivity analysis using a cutoff of 1.1 (vs. 0.8 in the main analysis) did not significantly affect our findings (Table S4).

**Table S4: Sensitivity analysis with higher seropositivity threshold**

| **Variable** | **Category** | **Model 1** | **Model 2** | **Model 3** |
| --- | --- | --- | --- | --- |
| **Age** | <18 | 2.27 (1.63-3.17) | 2.15 (1.55-2.98) | 2.18 (1.56-3.05) |
|  | 18-29 | 1.52 (1.07-2.16) | 1.46 (1.03-2.08) | 1.45 (1.02-2.07) |
|  | 30-44 | (Ref) | (Ref) | (Ref) |
|  | 45-59 | 0.79 (0.53-1.17) | 0.82 (0.55-1.21) | 0.81 (0.54-1.2) |
|  | >=60 | 1.22 (0.72-2.06) | 1.33 (0.79-2.24) | 1.31 (0.77-2.2) |
| **Gender** | Male | (Ref) | (Ref) | (Ref) |
|  | Female | 1.55 (1.22-1.98) | 1.53 (1.2-1.95) | 1.54 (1.2-1.96) |
| **Children in home (Y/N)** |  | 1.31 (0.93-1.84) |  | 0.9 (0.61-1.32) |
| **Total residents** |  |  | 1.22 (1.11-1.34) | 1.24 (1.11-1.38) |
| ***AIC*** |  | *2617* | *2602* | *2603* |
| ***ICC*** |  | *0.409* | *0.399* | *0.399* |

SARS-CoV-2 serology results were available for all residents of 403 households, accounting for 917 individuals (45.1% of our total study population). In order to assess the robustness of our findings, we compared the characteristics of households with and without complete serological data for all residents. Panels A and B in Figure S3 show the distribution of total number of residents and number of seropositive individuals per household. Odds ratios in univariable analyses did not differ significantly between the whole study population and the subset of households with complete serological data (Figure S4).

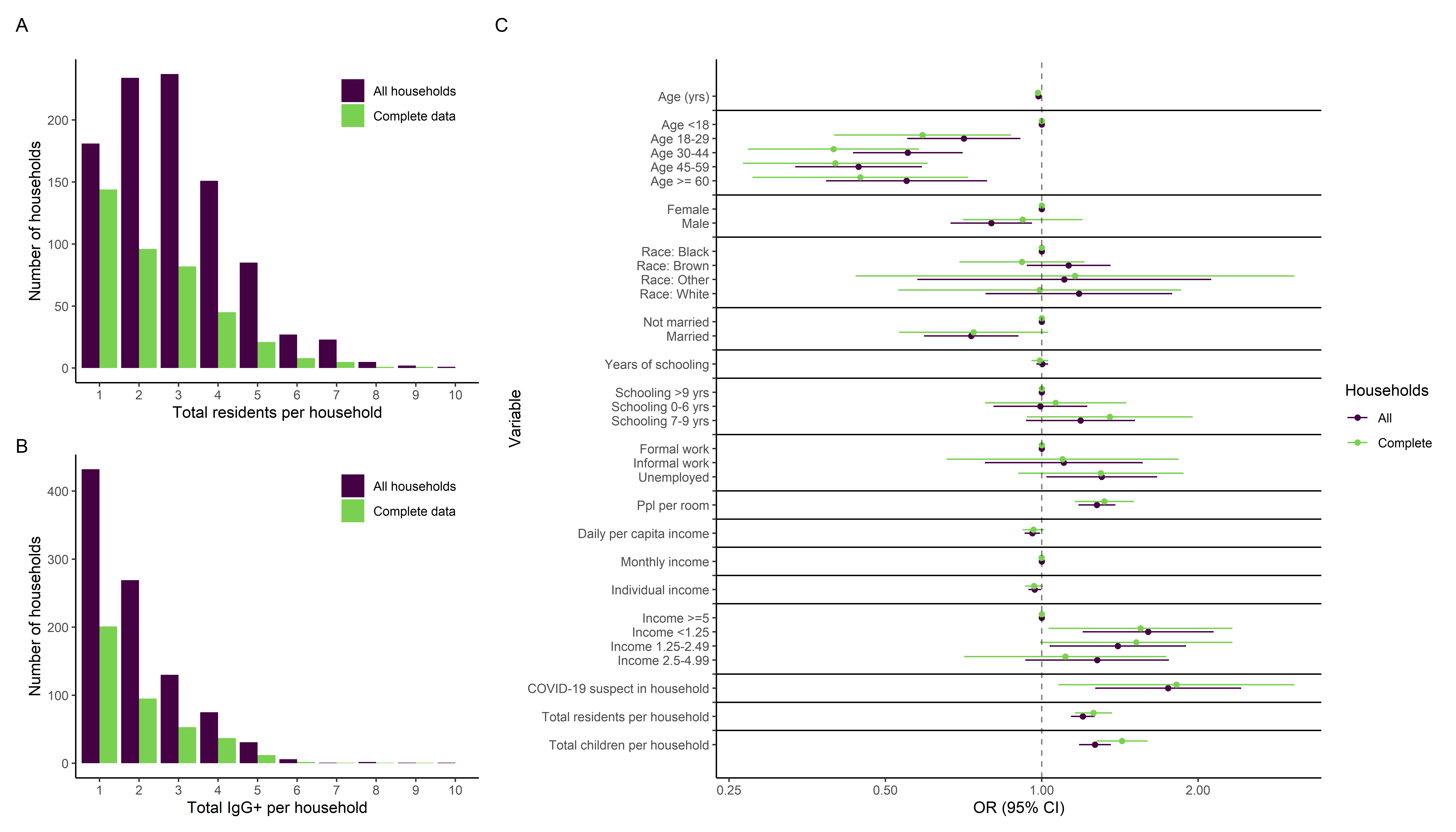

**Figure S4**: Comparison of households with complete serological data and overall study population. Distribution of total number of residents (A) and seropositive individuals (B) in each household. Panel C shows univariable odds ratios for all households compared to those with serological data for all residents (“complete”).

We repeated our primary analyses, using only data from households with complete serological data. The distribution of age and association with SARS-CoV-2 seropositivity was similar among the subset of households with complete serological data (Figure S5). The associations between gender, household composition, income, and SARS-CoV-2 seropositivity remained similar in analyses including only households with complete serological data (Figure S6).

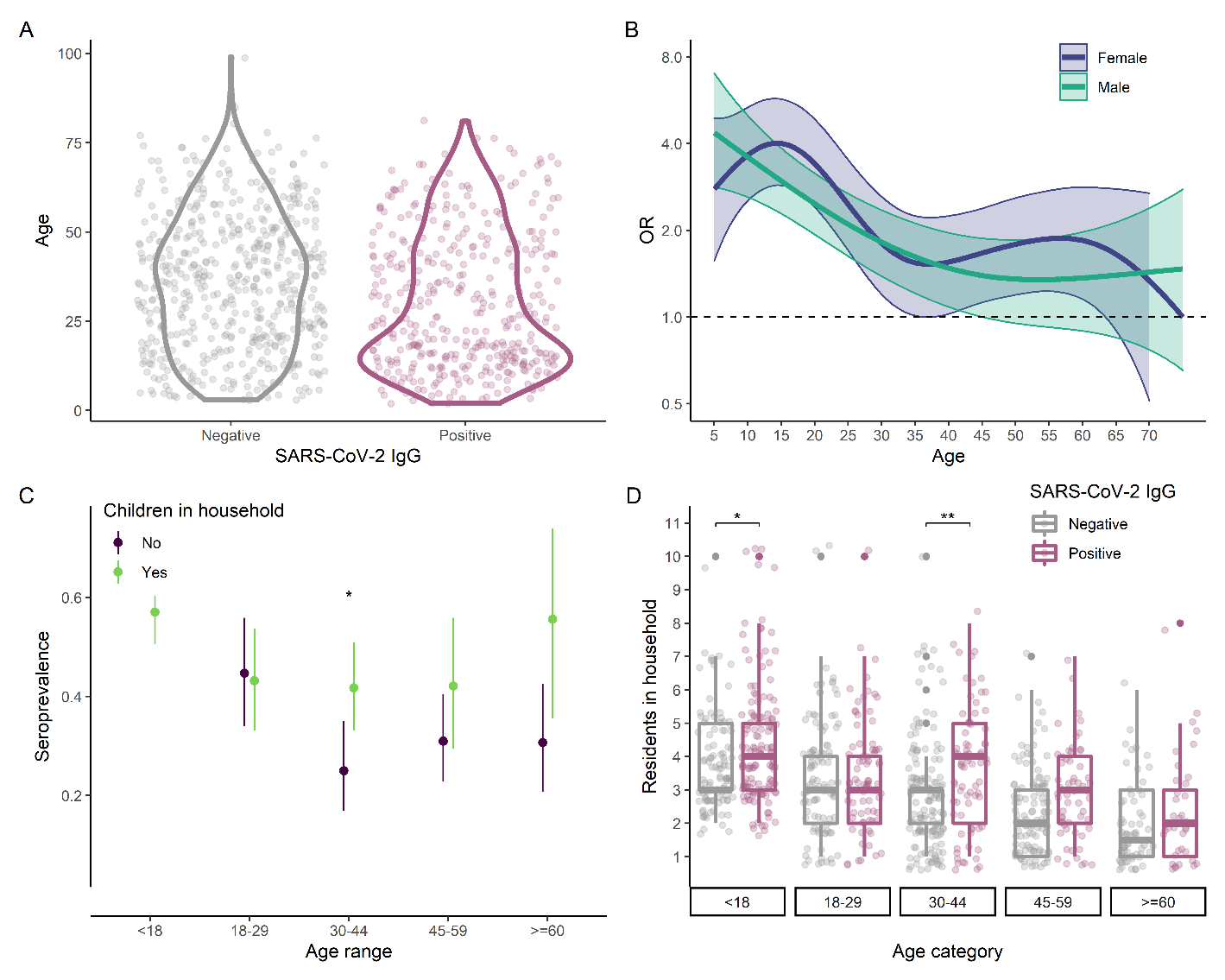
**Figure S5**: Sensitivity analysis on effect of age, among households with complete serological data. A: Distribution of age between SARS-CoV-2 seronegative and seropositive individuals. B: Relationship between age and SARS-CoV-2 seropositivity by gender, as estimated using a generalized additive model. C: Seroprevalence by age, with and without presence of children in the household. D: Comparison of household size and age between SARS-CoV-2 seronegative and seropositive individuals. Asterisks indicate statistically significant differences (Bonferroni-adjusted p<0.05: *; <0.01: **; <0.001: ***).

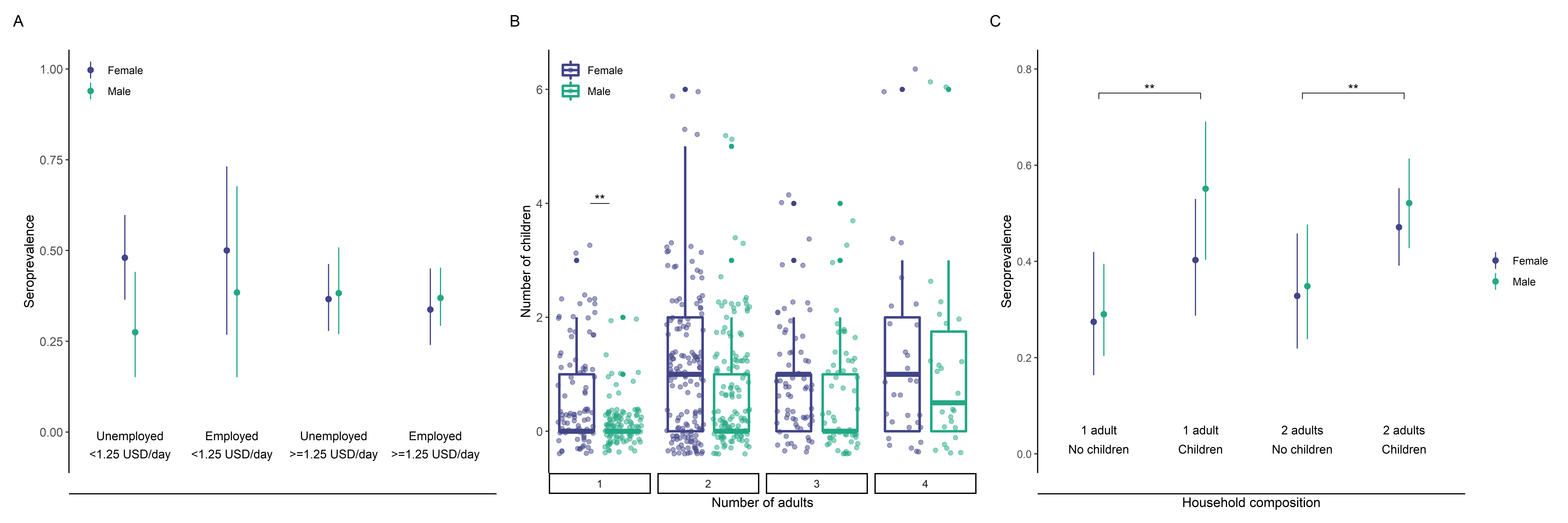
**Figure S6**: Sensitivity analysis on the effects of gender, income, and household composition, among households with complete serological data. A: Seroprevalence by daily per capita income, employment and gender. B: Comparison of household composition, assessed by number of adults and children, by gender. C: Variation in seroprevalence by household composition. Asterisks indicate statistically significant differences (Bonferroni-adjusted p<0.05: *; <0.01: **; <0.001: ***).

Because age was strongly associated with income, we did not include both variables simultaneously in our multilevel multivariable regression model. An alternative model including daily per capita income rather than age yielded similar findings: the increased seroprevalence in households with children was mediated by increased household size (Table S5).

**Table S5: Sensitivity analysis with income**

|  |  | **Model 3** | **Model 4** | **Model 5** |
| --- | --- | --- | --- | --- |
| **Variable** | **Category** |  |  |  |
| **Income** | <1.25 | 1.51 (1-2.28) | 1.29 (0.85-1.97) | 1.26 (0.82-1.92) |
|  | 1.25-2.49 | 1.24 (0.8-1.93) | 1.08 (0.7-1.68) | 1.06 (0.68-1.65) |
|  | 2.5-4.99 | 1.22 (0.78-1.91) | 1.16 (0.74-1.8) | 1.14 (0.73-1.77) |
|  | ≥5 | (Ref) | (Ref) | (Ref) |
| **Gender** | Male | (Ref) | (Ref) | (Ref) |
|  | Female | 1.3 (1.02-1.65) | 1.31 (1.03-1.67) | 1.3 (1.02-1.66) |
| **Children in home (Y/N)** |  | 1.71 (1.25-2.34) |  | 1.23 (0.86-1.75) |
| **Total residents** |  |  | 1.25 (1.14-1.37) | 1.22 (1.1-1.35) |
| ***AIC*** |  | *2358* | *2343* | *2344* |
| ***ICC*** |  | *0.329* | *0.319* | *0.319* |
